# The impact of COVID-19 non-pharmaceutical interventions on future respiratory syncytial virus transmission in South Africa

**DOI:** 10.1101/2022.03.12.22271872

**Authors:** Samantha Bents, Cécile Viboud, Bryan Grenfell, Alexandra Hogan, Stefano Tempia, Anne von Gottberg, Jocelyn Moyes, Sibongile Walaza, Cheryl Cohen, Rachel Baker

**Affiliations:** Department of Ecology and Evolutionary Biology, Princeton University, Princeton, NJ, USA; Fogarty International Center, National Institutes of Health, Bethesda, Maryland, United States; School of Population Health, University of New South Wales, Sydney, New South Wales, Australia; Centre for Respiratory Diseases and Meningitis, National Institute for Communicable Diseases of the National Health Laboratory Service, Johannesburg, South Africa; School of Public Health, Faculty of Health Sciences, University of Witwatersrand, Johannesburg, South Africa; Princeton Environmental Institute, Princeton University, Princeton, NJ, USA; School of Pathology, Faculty of Health Sciences, University of Witwatersrand, Johannesburg, South Africa; Department of Pathology, Faculty of Health Sciences, University of Cape Town, Cape Town, South Africa

## Abstract

In response to the COVID-19 pandemic, the South African government employed various nonpharmaceutical interventions (NPIs) in order to reduce the spread of SARS-CoV-2. In addition to mitigating transmission of SARS-CoV-2, these public health measures have also functioned in slowing the spread of other endemic respiratory pathogens. Surveillance data from South Africa indicates low circulation of respiratory syncytial virus (RSV) throughout the 2020-2021 Southern Hemisphere winter seasons. Here we fit age-structured epidemiological models to national surveillance data to predict the 2022 RSV outbreak following two suppressed seasons. We project a 32% increase in the peak number of monthly hospitalizations among infants ≤ 2 years, with older infants (6-23 month olds) experiencing a larger portion of severe disease burden than typical. Our results suggest that hospital system readiness should be prepared for an intense RSV season in early 2022.

## Introduction

RSV is the most common cause of lower respiratory infections and is responsible for an estimated 5% of under-five mortality globally [1]. In most countries, RSV circulation has been disrupted due to COVID-19, yet the consequences of two years of disrupted transmission patterns on future RSV seasons remain debated. Since the start of the COVID-19 pandemic, South Africa has experienced four large waves of SARS-CoV-2 infection driven by different variants, with varying levels of control. A 5-level alert system was put in place to quantify the strength of nonpharmaceutical interventions (NPIs) to mitigate COVID-19 in March of 2020 [2]. NPIs were instituted based on the COVID-19 national risk, and included social distancing, travel bans, school closures, mask wearing, and other government measures used to slow COVID-19 transmission. These NPIs had major effects on behavior and repressed circulation of a range of communicable respiratory diseases, with record low RSV and influenza seasons reported in 2020 [2, 3]. As NPIs were relaxed in the spring of 2020, RSV made a late out-of-season resurgence that persisted into 2021. Despite this rebound, RSV transmission has remained historically low [2]. Understanding the potential hospital burden of RSV in 2022 following two suppressed seasons, and possible shifts in the demography of affected children, is a key public health question.

Several birth cohort studies have shed light on RSV transmission patterns and predictors of disease severity [4, 5]. Almost all infants are infected by the age of 2 years, but severe infections occur primarily in infants ≤ 6 months of age [6, 7]. Infections among infants are primarily driven by transmission from older siblings in high-contact settings, such as schools and childcare centers [8]. Children may be reinfected as RSV immunity wanes on short time scales, but secondary infections are unlikely to result in hospitalization [6]. It is uncertain whether milder infections at older ages are a product of more mature immune systems or immune protection from prior infections [9]. Regardless, preventing and treating severe primary infection to RSV in the first two years of life has been the primary focus of disease mitigation strategies [10]. The distinct age-structure of RSV burden may have implications for hospitalizations during future outbreaks; this is because the implementation of NPIs throughout the pandemic reduced endemic pathogen circulation globally, enabling the buildup of fully naive individuals ≤ 2 years old [11]. This poses the risk of a large and potentially severe RSV outbreak as NPIs are relaxed and a large number of infants are exposed to RSV for the first time.

The timing of a potential impending outbreak is important to consider in addition to magnitude. RSV circulation is driven in part by low specific humidity and typically occurs in wintertime [12]. Unlike in most other temperate settings, RSV peaks in the autumn between February-June before the influenza season in South Africa [13]. If NPIs are relaxed throughout the time period of typical seasonal transmission, climate drivers could further impact the magnitude and timing of an outbreak.

We use an epidemiological model to predict the age-structure, timing, and magnitude of a 2022 RSV outbreak in South Africa. We consider the underlying seasonality and buildup of susceptible infants over the last two years and project the hospital burden among infants ≤ 2 years of age. We explore the public health implications of the model projections for a lower middle-income country like South Africa.

## Data

This study uses facility-based data collected through the Severe Acute Respiratory Illness (SARI) and Influenza-like Illness (ILI) Surveillance Programmes from January 2015-August 2021 (weeks 1-33). ILI and SARI surveillance is conducted among outpatients in 5 public health clinics and inpatients in 7 hospitals respectively. The hospitals and clinics span 5 provinces: Western Cape, North West, Gauteng, KwaZulu-Natal, and Mpumalanga. We used weekly numbers of RSV-positive cases disaggregated by month for children ≤ 5 years. Respiratory specimens were collected among enrolled patients meeting the case definition using nasopharyngeal aspirates or nasopharyngeal and oropharyngeal swabs. Positive cases were confirmed using real-time reverse transcription PCR (rtRT-PCR) [2].

## Methods

We use an age-structured model originally described by Hogan et al. and adapt it to South Africa age-structured case observations [10]. The full SEIRS model is given by:

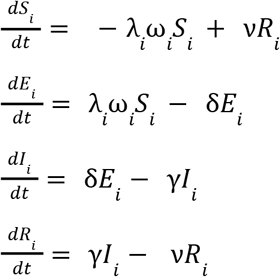

where *S* is the susceptible population; *E* is recovered; *I* is infected; *R* is recovered, and *i* represents the given age cohort. 1/γ is the infectious period set to 9 days and 1/δ is the latent period set to 4 days. We vary the immunity period 1/ν (see Table 1 for ranges) and set it to 150 days in the main analysis, which is the value that minimized the mean squared errors between observations and predictions in the pre-pandemic period. We define the pre-pandemic period as the period from January 2015-December 2019, and the pandemic period from January 2020-August 2021. ω_*j*_ represents reduced infectiousness in children older than 10 years.

**Table 1:** Model Parameters and parameter ranges. Fixed model parameters and the range of estimates considered for each parameter fit to surveillance data.

| Parameter | Definition | Range of Estimates | Value |
| --- | --- | --- | --- |
| $\nu$ | Duration of immunity (days) | 60-720 | 150 |
| $\beta_1$ | Amplitude of seasonal forcing | .1-.3 | .3 |
| $\gamma$ | Infectious period (days) | Fixed [10] | 9 |
| $\delta$ | Latent period (days) | Fixed [10] | 4 |
| $\phi$ | Phase shift | 0-2π | 2.0 |
| $\beta_0$ | Transmission coefficient | Fixed [10] | .03 |
| $h_1, h_2, h_3, h_4$ | Hospitalization scaling factors | 0-.5 | .41, .10, .05, .02 |

The function used to capture the RSV transmission rate on age class *i* is given by:

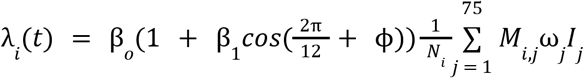

Here *λ*_*i*_(*t*) is the monthly transmission rate and *t* is time in months, *β*_1_ and Φ represent the amplitude and phase respectively that capture seasonality in transmission in South Africa. *M*_*i,j*_ describes the contact matrix between individuals in age group *j* and *i* between 0-75 years of age, considering the age of children ≤ 5 years by one month bands and persons aged 5-75 by 5 year bands. We use an expanded version of the contact matrix originally described by Mossong et al., which describes population mixing patterns for eight European countries [14]. We fit the seasonal coefficients *β*_1_ and Φ to pre-pandemic observations [Appendix I]. *β*_0_ is the transmission coefficient fixed at .03 for all locations, based on prior work [10].

Incidence is scaled to SARI surveillance data by fitting hospitalization parameters for infants 0-2 months, 3-5 months, 6-11 months, and 12-23 months of age. We determine South Africa-specific hospitalization scaling factors *h*_*1*,_ *h*_*2*,_ *h*_*3*,_ *h*_*4*_ to be .41, .10, .05, and .02 for the four respective age groups. We capture the age structure of severe infection in South Africa by scaling model incidence to the average number of hospitalizations in each of the four key age groups calculated during 2015-2019.

Finally, we use data from the pandemic period to estimate how control measures implemented to prevent the spread of COVID-19 have affected RSV transmission. We measure strength of the control periods with c, the percent reduction in transmission λ_*i*_(*t*). The adjusted equation then can be described as:

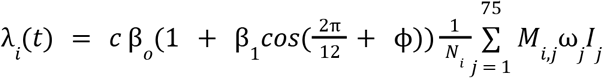

where 0 < *c* < 1. We fit two control periods to describe the two intervals when the South African government instituted the highest lockdown levels (levels 4 or 5). We choose to only consider lockdown levels for two periods which coincided with the extended closure of schools, a measure which has been found to directly reduce the circulation of childhood infections such as RSV [15]. These periods of intense lockdown were instituted to control the first and third COVID-19 waves which occurred in early 2020 and mid-2021. We determine *c* for both lockdown periods by minimizing the mean squared error between model predictions and the observed hospitalizations in the pandemic period.

After calibrating the model to pre-pandemic data and estimating the impact of 2020-2021 interventions, we let the model run during 2022-2027 to project RSV trajectory in future years. In the main analysis, we consider that interventions are fully relaxed in 2022 and beyond. Based on model projections, we estimate the size of the 2022 peak and total outbreak size relative to pre-pandemic years, and the age distribution of hospitalizations in the 2022 outbreak. We also evaluate the periodicity and timing of outbreaks after 2022.

In sensitivity analyses, we consider a range of future NPI scenarios given the possibility of persistent COVID-19 circulation [Appendix IV]. We explore scenarios representing different strengths of NPIs, corresponding to 20% and 35% reductions in transmission (c = .20, and .35), representing low and high levels of NPI. We also vary the length of NPIs between 6, 12, and 36 months. This allows us to compare six hypothetical scenarios ranging from low levels of persistent behavioral changes to full lockdowns in response to potential new variants.

## Results

### RSV patterns in 2020-2021 surveillance data

The NPIs implemented to control COVID-19 caused disrupted RSV transmission patterns as shown by Fig. 1 (top). RSV was almost completely suppressed during the typical transmission period in 2020 but made a resurgence in August-December of 2020, coinciding with the relaxation of NPIs. The strong, out of season outbreak recorded approximately 70% of the number of hospitalizations in infants ≤ 2 years old relative to 2019. Transmission continued into 2021 following normal seasonal patterns but was reduced as a result of the implementation of stricter NPIs to control the third COVID-19 wave in South Africa (Beta variant). Age-structured surveillance data also provides evidence that the disruption in transmission affected the age distribution of severe infections compared to pre-pandemic years. Fig. 1 (bottom) shows the proportion of hospitalizations within four high-risk age groups (0-2 months, 3-5 months, 6-11 months, 12-23 months) in 2020-2021 compared to a five year pre-pandemic average.

**Figure 1:**
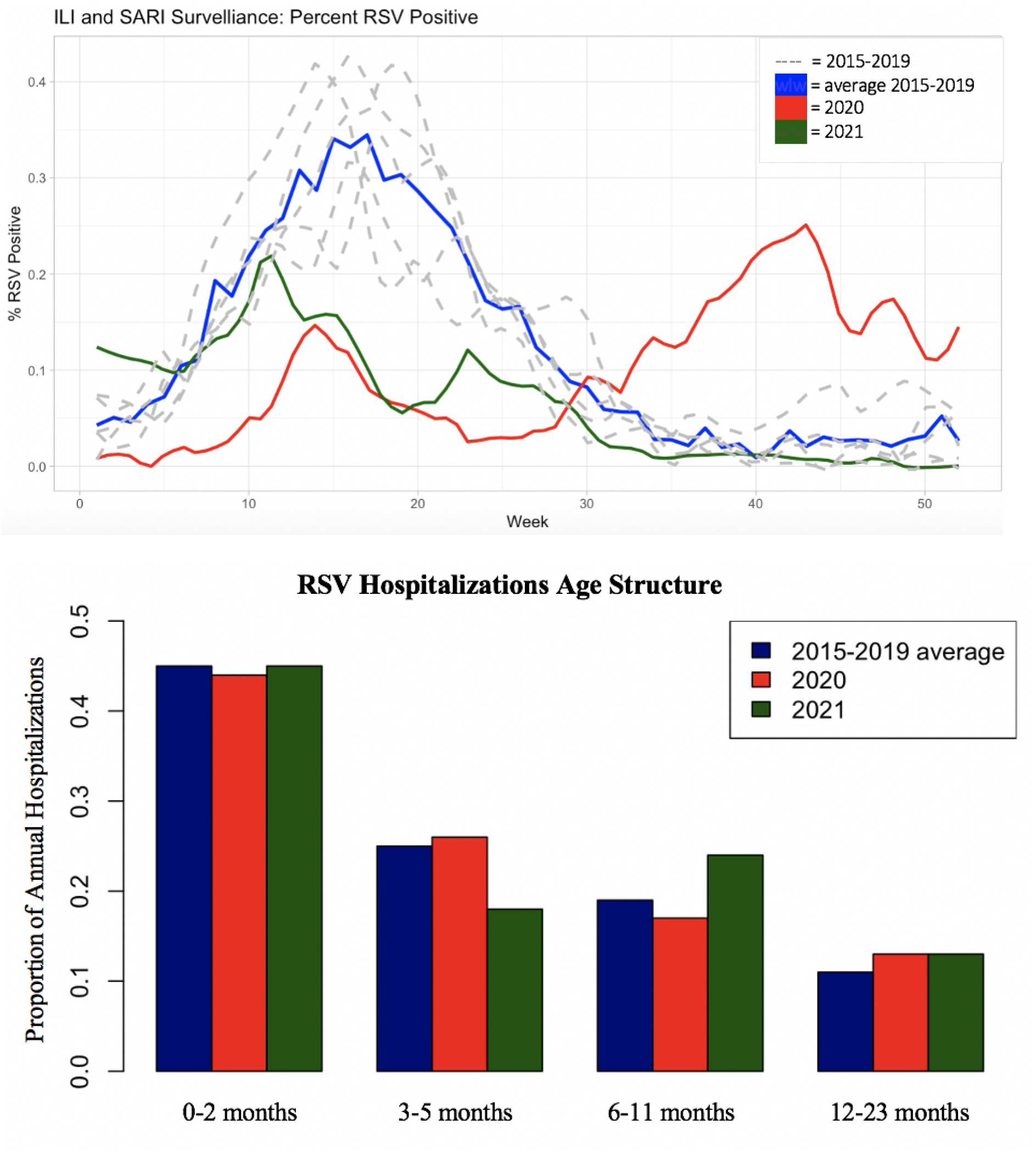
Viral activity and age distribution of RSV in 2015-2021, South Africa. Top: Surveillance Time Series for RSV in South Africa. The percent positive RSV tests for the 2020 season (red) and 2021 season (green) compared to the average for five preceding seasons, which follows the typical seasonal transmission pattern (blue). Percent positive for individual years 2015-2019 (dashed gray) show typical seasonal variability. **Bottom: Age shift in RSV surveillance data coinciding with the COVID-19 pandemic**. The proportion of annual hospitalizations observed in 0-2 months, 3-5 months, 6-11 months, and 12-23 months age groups in 2020-2021 compared to the average for five preceding seasons. While the proportion of annual hospitalizations in the 0-2 and 12-23 months age groups remains consistent, we observe a shift from 3-5 months to 6-11 months.

**Figure 2:**
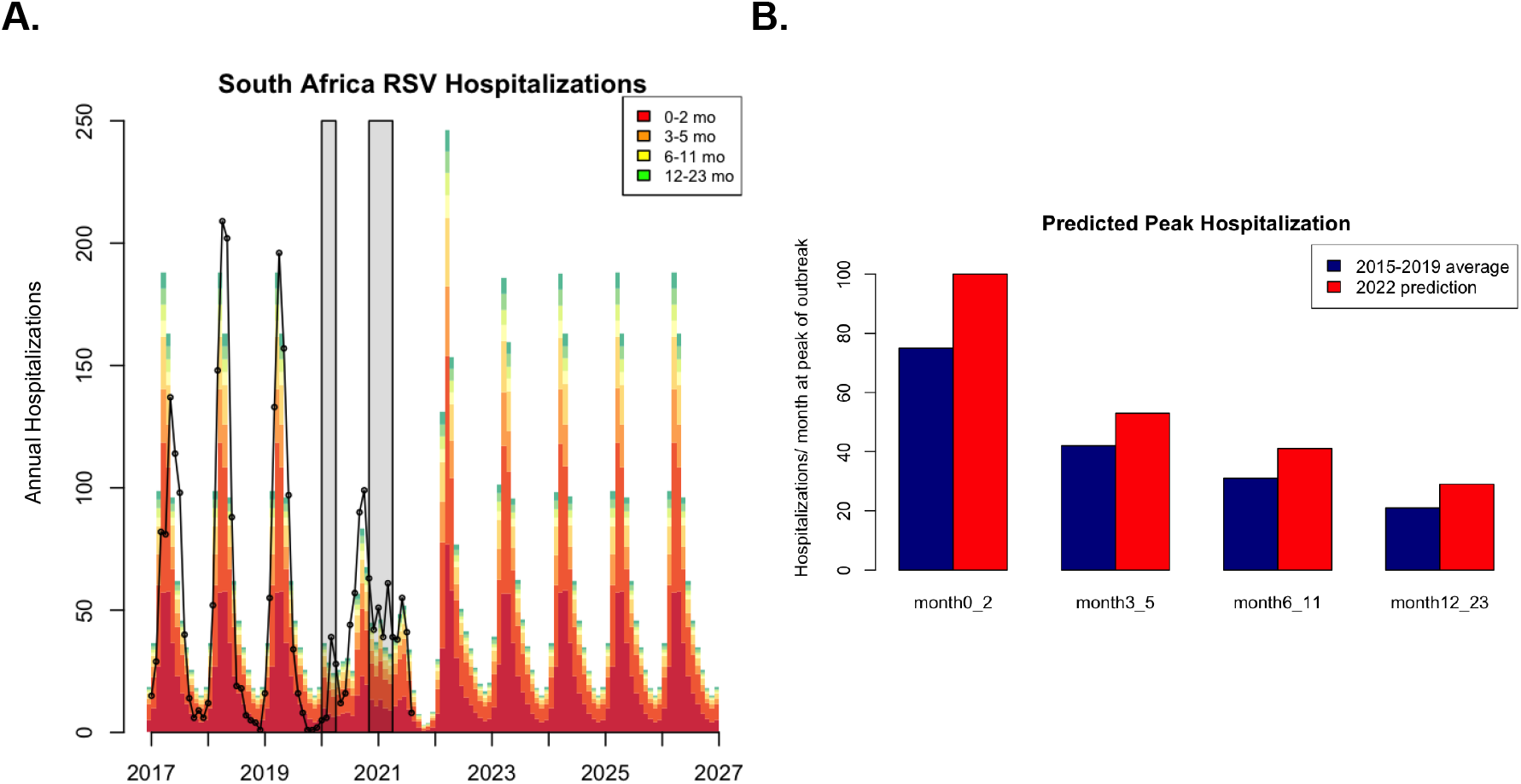
Model Predictions. A) Age-structured model of the post-NPI RSV resurgence in South Africa fit to national case data (black dots). Gray bars indicate the first and second control periods. B) Predicted 2022 peak hospitalizations by age group compared to the average recorded for five preceding seasons (2015-2019). We see a large increase in all older infant classes ≤ 2 years at the peak of the outbreak.

### RSV model calibration to data

In order to understand the magnitude and timing of potential RSV resurgence in 2022 following two irregular RSV seasons, we apply the age-structured epidemiological model to project scenarios for future disease burden. We input control periods into the model in line with the timing of two strict lockdowns implemented by the government to control the spread of COVID-19, which included school closure. We estimate a 47% and 37% decrease in RSV transmission for the first and second lockdown periods respectively.

Of particular interest for public health is the quantification of the peak magnitude and intensity of RSV, which is important for hospital surge capacity, especially in neonatal and perinatal wards. The model predicts an irregularly intense peak in 2022, suggesting the outbreak will occur more rapidly than in previous years. According to our projections, circulation builds quickly, and the outbreak reaches peak incidence in April 2022. At the peak of the 2022 outbreak, we predict there will be a 32% increase in the number of monthly hospitalizations compared to the average recorded for five preceding seasons (2015-2019), with the largest percent increase being among older age groups. Our projections show a 26, 31, and 36% increase in peak hospitalizations for the 3-5, 6-11, and 12-23 month age groups respectively.

While the hospital burden among 0-2 months old remains considerable, we anticipate that the largest proportional age shift in hospitalizations will be among 6-23 month olds. Infants > 2 months olds are typically less likely to experience severe infection, but reduced RSV circulation in the two preceding RSV seasons increases the number of infants susceptible to primary infection in the 6-23 month age group [6]. The model also projects that the disruption to RSV circulation will be short-lived and typical RSV seasons will resume by 2023 assuming further school closures are not implemented.

We additionally consider the potential for NPIs of different lengths and intensities ranging over the next 1-3 years [Appendix IV]. We find that low levels of NPIs (β reduced by < 20%) have a negligible effect on model predictions, with projections still showing an intense 2022 outbreak. At stricter levels of NPI (β reduced by > 35%) we determine the possibility of an intense but smaller 2022 RSV season, further increasing the buildup of susceptible individuals and leading to a more intense 2023 outbreak relative to 2022 predictions. As the length of the NPI period increases to 3 years, we find that the likelihood of a large outbreak occurring when measures are lifted decreases and transmission returns to typical dynamics more quickly as a result of transmission persisting during the NPI period.

## Discussion

We anticipate an intense RSV outbreak in 2022, with a peak projected to occur in April 2022 and a 32% increase in magnitude relative to pre-pandemic seasons. Hospitalization burden at the peak of the season will be higher than in pre-pandemic seasons in all age groups under 2 years, but the largest proportional increase will be in 6-23 month olds. The projected age shift aligns with observed patterns during the pandemic in South Africa, where almost 40% of annual hospitalizations in 2021 were within the 6-23 month age group, representing a 20% increase compared to pre-pandemic data. The trend towards older ages suggests that RSV severity is considerably influenced by prior immunity.

Due to the possibility of a large RSV outbreak, alongside the possible resurgence of other endemic pathogens that have been repressed during COVID-19, there are various key public health questions to consider [3]. We predicted that older age groups (6-23 months) will experience the largest proportional age shift in hospitalizations, driven by an increase in the population susceptible to primary RSV infection. Hospitals may consider preparing for a larger number of babies experiencing severe primary infections at older ages than typical. We recommend the healthcare system prepares for a condensed RSV season, with an earlier than usual peak number of RSV-related hospitalizations in early April 2022. In response to this possibility, hospitals may consider increasing the availability of prophylaxis to high-risk infants in order to prevent severe infection and hospitalization, although there is limited use of these expensive treatments in routine in South Africa. More broadly, countries around the world such as France, Japan, and Australia, among others, have reported large, out of season RSV outbreaks following the lift of COVID-19 NPIs, reflecting a global pattern in the nature of RSV resurgence in response to pandemic-related public health measures [8, 16, 17, 18].

These findings are important to consider in the context of a lower middle-income country (LMIC), in which diagnostic tests may be limited and infants tend to be at higher risk for developing severe symptoms upon RSV infection [19, 20]. Given the high cost of current prophylaxis treatments such as Palivizumab, the potential hospital burden of an outbreak occurring over a condensed period of time may be considerable [6, 21]. Hospitals may also be dually impacted by the downstream effects of reduced health seeking throughout the pandemic. From March 2020-September 2021, total hospital admissions dropped by 60% in South Africa, indicating that non-COVID-19 illnesses have been both reduced and gone increasingly untreated in recent years [3]. As general health seeking returns to pre-pandemic levels, there may be a surge in individuals needing hospital care [3]. Persistent COVID-19 circulation may additionally pose a burden on the healthcare system. Hospitals in South Africa have reported an increase in the number of COVID-19 hospitalizations among young children as a result of the Omicron variant, with the number of pediatric hospitalizations uncharacteristically surpassing adult hospitalizations in some locations [22]. This further highlights the importance of preparing the healthcare system for an intense RSV outbreak.

RSV circulation in 2022 will be dependent on the NPIs that are implemented by the South African government. Without strong NPIs in place throughout the typical RSV season, South Africa is likely to see a larger than average RSV outbreak in 2022 as modeled in this study. Given that no additional NPI measures were put in place in response to the rise of the COVID-19 Omicron variant in late 2021 and NPIs continue to remain relaxed in early 2022, it is unlikely that RSV circulation will be suppressed for a third year. Masking and other behavioral changes however could continue at low levels and have a moderate impact on transmission. Most educational institutions have maintained mask mandates into 2022, potentially reducing transmissibility among children in high-contact settings [23]. This study focuses primarily on modeling the impact of lockdowns including school closures on RSV transmission, but implicitly captures the effect of masking at school in fitting the model to surveillance data from the pandemic period. Our sensitivity analyses suggest that persistence of low levels of NPIs, consistent with masking, would have a negligible effect on model projections and would not substantially attenuate the intensity of the 2022 outbreak. Future studies however should explore the specific role of masking in preventing the transmission of RSV in educational settings.

There are several caveats to our model predictions. First, we use an age-contact structure derived from European data in our model; although some local data exist from South Africa, the fit of the model using these data was not as good (not shown). This study uses a short duration of RSV immunity (150 days), on the basis of it producing an optimal model fit to pre-pandemic data, as information on this parameter is scarce. In order to test the validity of this parameter, immunity was varied between 2-24 months (Appendix II). For immunity lasting beyond 150 days, the model overestimated the length of the typical RSV season in South Africa. Further, the model used does not distinguish between primary and secondary infections in relation to transmissibility and severity. We assume instead that age is the primary predictor of whether an infection will result in hospitalization. By fitting the age-structured model to age-structured observations, we show that the model predicts the distribution of hospitalizations among the studied age groups with confidence. We additionally conduct a sensitivity analysis by fitting a time-series SIR (TSIR) model which assumes a fully immunizing infection (lifelong immunity) to observed data [24]. Using the TSIR model we predict that the outbreak will occur earlier than typical and be about 3 times larger than average at the peak of infection [Appendix III]. Comparison between the two models suggest that the age-structured short-term immunity SEIRS model may trend towards underestimating potential hospital burden; hence projections presented in our main analyses should be deemed conservative. Overall, the two modeling frameworks, TSIR and SEIRS, bracket the space of plausible RSV trajectories; it is notable that both consistently support the possibility of an intense outbreak in 2022.

## Conclusion

This paper explores the indirect impacts of COVID-19 NPIs on RSV transmission in South Africa, demonstrating the importance of studying the public health implications of suppressed seasonal infections. It represents the first age stratified RSV analysis completed in an LMIC country following the COVID-19 pandemic and highlights the potential need for increased hospital readiness to address a large and intense RSV outbreak in 2022.

## Data Availability

Data is available online through the National Institute for Communicable Diseases at https://www.nicd.ac.za/.

## References

1. Wang, H. et al. Global, regional, and national life expectancy, all-cause mortality, and cause-specific mortality for 249 causes of death, 1980–2015: a systematic analysis for the Global Burden of Disease Study 2015. The Lancet 388, 1459–1544x (2016).

2. Tempia, S. et al. Decline of influenza and respiratory syncytial virus detection in facility-based surveillance during the COVID-19 pandemic, South Africa, January to October 2020. Eurosurveillance 26, (2021).

3. Perofsky, A. C. et al. The direct and indirect effects of the COVID-19 pandemic on private healthcare utilization in South Africa, March 2020 – September 2021. Clinical Infectious Diseases ciac055 (2022) doi:10.1093/cid/ciac055

4. Ohuma, E. O. et al. The Natural History of Respiratory Syncytial Virus in a Birth Cohort: The Influence of Age and Previous Infection on Reinfection and Disease. American Journal of Epidemiology 176, 794–802 (2012).

5. Hardelid, P., Verfuerden, M., McMenamin, J., Smyth, R. L. & Gilbert, R. The contribution of child, family and health service factors to respiratory syncytial virus (RSV) hospital admissions in the first 3 years of life: birth cohort study in Scotland, 2009 to 2015. Eurosurveillance 24, (2019).

6. Sommer, C. Risk Factors for Severe Respiratory Syncytial Virus Lower Respiratory Tract Infection. TOMICROJ 5, 144–154 (2011).

7. Staadegaard, L. et al. The Global Epidemiology of RSV in Community and Hospitalized Care: Findings From 15 Countries. Open Forum Infectious Diseases 8, ofab159 (2021).

8. van Summeren, J. et al. Low levels of respiratory syncytial virus activity in Europe during the 2020/21 season: what can we expect in the coming summer and autumn/winter? Eurosurveillance 26, (2021).

9. Rodríguez, D. A. et al. Predictors of severity and mortality in children hospitalized with respiratory syncytial virus infection in a tropical region: Epidemiology of RSV in a Tropical Region. Pediatr Pulmonol. 49, 269–276 (2014).

10. Hogan, A. B. et al. Potential impact of a maternal vaccine for RSV: A mathematical modelling study. Vaccine 35, 6172–6179 (2017).

11. Baker, R. E. et al. The impact of COVID-19 nonpharmaceutical interventions on the future dynamics of endemic infections. Proc Natl Acad Sci USA 117, 30547–30553 (2020).

12. Baker, R. E. et al. Epidemic dynamics of respiratory syncytial virus in current and future climates. Nat Commun 10, 5512 (2019).

13. Kyeyagalire, R. et al. Hospitalizations associated with influenza and respiratory syncytial virus among patients attending a network of private hospitals in South Africa, 2007–2012. BMC Infect Dis 14, 694 (2014).

14. Mossong, J. et al. Social Contacts and Mixing Patterns Relevant to the Spread of Infectious Diseases. PLoS Med 5, e74 (2008).

15. Zheng, Z., Pitzer, V. E., Warren, J. L. & Weinberger, D. M. Community factors associated with local epidemic timing of respiratory syncytial virus: A spatiotemporal modeling study. Sci. Adv. 7, eabd6421 (2021).

16. Casalegno, J.-S. et al. Characteristics of the delayed respiratory syncytial virus epidemic, 2020/2021, Rhône Loire, France. Eurosurveillance 26, (2021)

17. Eden, J.-S. et al. Off-season RSV epidemics in Australia after easing of COVID-19 restrictions. http://medrxiv.org/lookup/doi/10.1101/2021.07.21.21260810 (2021) doi:10.1101/2021.07.21.21260810.

18. Ujiie, M., Tsuzuki, S., Nakamoto, T. & Iwamoto, N. Resurgence of Respiratory Syncytial Virus Infections during COVID-19 Pandemic, Tokyo, Japan. Emerg. Infect. Dis. 27, 2969–2970 (2021).

19. Sonego, M., Pellegrin, M. C., Becker, G. & Lazzerini, M. Risk Factors for Mortality from Acute Lower Respiratory Infections (ALRI) in Children under Five Years of Age in Low and Middle-Income Countries: A Systematic Review and Meta-Analysis of Observational Studies. PLoS ONE 10, e0116380 (2015).

20. Fleming, K. A. et al. The Lancet Commission on diagnostics: transforming access to diagnostics. The Lancet 398, 1997–2050 (2021).

21. Ginsberg, G. M., Somekh, E. & Schlesinger, Y. Should we use Palivizumab immunoprophylaxis for infants against respiratory syncytial virus? – a cost-utility analysis. Isr J Health Policy Res 7, 63 (2018).

22. Cloete, J. et al. Rapid rise in paediatric COVID-19 hospitalisations during the early stages of the Omicron wave, Tshwane District, South Africa. http://medrxiv.org/lookup/doi/10.1101/2021.12.21.21268108 (2021) doi:10.1101/2021.12.21.21268108.

23. South African Government Department of Health. Face Masks. (2022). https://www.sahealth.sa.gov

24. Becker, A. D. & Grenfell, B. T. tsiR: An R package for time-series Susceptible-Infected-Recovered models of epidemics. PLoS ONE 12, e0185528 (2017).

